# Anopheles mosquito exposure is associated with age, gender and bed net use in areas in Uganda experiencing varying malaria transmission intensity

**DOI:** 10.1101/2024.12.12.24318757

**Authors:** Sara Lynn Blanken, Maxwell Kilama, Jordache Ramjith, Alex K. Musiime, Kjerstin Lanke, Daniel Ayo, Kristiaan Huijbers, Tom Hofste, Melissa Conrad, Paul Krezanoski, Grant Dorsey, Moses R. Kamya, Emmanuel Arinaitwe, Teun Bousema

## Abstract

**Objectives:** The number of *Anopheles* mosquito bites a person receives determines the risk of acquiring malaria and the likelihood of transmitting infections to mosquitoes. We assessed heterogeneity in *Anopheles* biting and associated factors in two settings in Uganda with different endemicity.

**Methods:** *Plasmodium falciparum* parasites in blood-fed indoor caught *Anopheles* mosquitoes were quantified using qPCR targeting the Pf18S rRNA gene. Human DNA in dried blood spots from household occupants and mosquito blood meals was profiled using 15 short-tandem repeats (STRs) and analysed using a log-likelihood approach for matching of both single and multi-sourced blood meals and incomplete DNA profiles.

**Results:** The distribution of mosquito bites was non-random; school-age children (5-15 years) and adults (≥16 years) had a mosquito biting rate ratio (BRR) 1.76 (95%CI 1.27-2.44, P < 0.001) and 1.96 (95%CI 1.41-2.73, P < 0.0001) times that of children under 5 years, respectively. Biting rates were lower in bed net users (BRR: 0.80, 95%CI 0.65-0.99, P = 0.042), and higher in males (BRR: 1.30, 95%CI 1.01-1.66, P = 0.043) and individuals infected with *P. falciparum* (BRR: 1.42, 95%CI 1.03-1.96, P = 0.030), though the latter effect lost statistical significance in sensitivity analyses.

**Conclusions:** Adults and school-age children are at higher risk for receiving mosquito bites and this has implications for the relative importance of demographic populations to onward malaria transmission to mosquitoes.

**Highlights:**

- A novel statistical approach allowed us to match multi-sourced mosquito bloodmeals and partially digested mosquito bloodmeals to human individuals
- Participant age consistently related to mosquito biting: adults and school-age children had a mosquito biting rate ratio that was ∼2 and ∼1.8 times that of children below 5 years, respectively
- Men received more mosquito bites than women and self-reported bed net use was associated with lower mosquito exposure.
- Higher mosquito biting rates were observed in individuals infected with *Plasmodium falciparum*

## Introduction

Infectious diseases typically exhibit heterogeneous transmission [1]. Certain individuals or populations may experience a disproportionately higher number of infection events. In mosquito-borne diseases, differences in mosquito exposure can contribute substantially to variations in pathogen exposure. Whilst mosquito exposure on a household level can be measured through mosquito catches that estimate mosquito density, considerable variation in mosquito exposure may exist between individuals from the same household [2, 3]. Non-random human biting frequency patterns are observed across mosquito species including *Anopheles* [4–10], *Culex* [10] and *Aedes* [2, 3, 11, 12].

The likelihood of receiving a mosquito bite depends on both host attractiveness and availability. Host availability is determined by host defensive behaviour and the amount of time spent unprotected from, yet in proximity of, mosquitoes. Previous studies on *Anopheles* mosquitoes have highlighted marked variation in the number of bites experienced by different demographic populations with younger children being bitten less frequently than would be expected [13]. Differences in host availability could partially explain these observed age-patterns in mosquito biting frequency [4, 5, 11, 14]. Adults were bitten more by mosquitoes in a site with high bed-net usage compared to a site with low [4] or no bed-net usage [8], suggesting that children slept under bed nets when available (net-usage was not reported).

Other host specific traits have also been associated with increased attractiveness to mosquitoes. Previous work in humans [4, 12] and birds [15] suggest a positive association between the number of received mosquito bites and body size, which for humans is strongly associated with age. A study in Papua New Guinea reported increased *Anopheles* biting on male compared to female study participants [14], although this difference was not observed by others studying biting preferences in *Anopheles* [5, 9] or *Aedes* [12] mosquitoes. At both long [16]- and short [7]-distances, pregnant women appear to attract more *Anopheles* mosquitoes compared to non-pregnant women.

Moreover, previous work using an olfactometer observed increased attractiveness to *Anopheles gambiae* mosquitoes in Kenyan children carrying microscopic densities of sexual-stage malaria parasites (i.e. gametocytes) [17, 18]. This difference disappeared when parasites were cleared with anti-malarial treatment, suggesting that differences in attractiveness to malaria vectors may be partially driven by infection status [17]. This observation underlines the relevance of quantifying mosquito exposure to understand both what populations are at highest risk of acquiring a mosquito-borne infection and what populations are most likely to transmit their infection to mosquitoes.

Attempts to understand heterogeneity in mosquito biting frequency have mainly focused on matching human DNA in mosquito bloodmeals. A common approach herein is profiling panels of short tandem repeats (STRs) and comparing STR profiles within a human population to mosquito bloodmeals [4, 6, 8, 14]. Using the existing methods, matching STR profiles requires an exact, or near-exact overlap of bloodmeal and human STR profiles. This impairs the ability to match multi-source bloodmeals that may comprise up to 22% of mosquito blood meals [19] and to match incomplete STR profiles that can primarily occur in low-blood volume mosquitoes [8] or in those that have partially digested their blood meals [20, 21]. Recently, a novel approach using log-likelihood ratios to match STR profiles was developed [22]. This log-likelihood approach enables matching of incomplete and multi-source STR profiles.

Here, we assessed *Anopheles* mosquito exposure and human biting rates in two settings in Uganda with different endemicity. By using a novel STR genotyping log-likelihood approach [22], we matched single-source and multi-source bloodmeals, including those with incomplete STR profiles. Our findings provide detailed insights into mosquito biting heterogeneity across different endemicity settings and identify human age, gender and bed net use as independent factors associated with mosquito exposure.

## Materials and Methods

### Study sites

This study was conducted in two different sites in eastern-Uganda. The first study took place between 2011 and 2019 in Nagongera subcounty, Tororo district (referred to as ‘Nagongera’). Due to implementation of indoor residual spraying (IRS) and long lasting insecticide treated nets (LLINS), malaria transmission declined from an entomological inoculation rate (EIR) of 238 in 2013 to 0.43 infective bites per person year in 2019 [23, 24]. The second study described here was conducted between August 2020 and July 2022 in 3 parishes comprising an area that spans the border between Tororo and Busia districts (referred to as ‘Tororo-Busia’). One parish was located in Busia district; the other two in Tororo district of which one is adjacent to the border with Busia district. In 2020, following a change in IRS formulation, the annual EIR in Tororo district was 27.4 and 59.0 for the parishes close and far from the Busia border, respectively. Busia district borders Tororo district to the south and is a high endemicity site with a reported annual EIR of 108.2 [25]. In both districts, LLINs were distributed universally in 2013, 2017 and 2020. IRS was implemented in Tororo district in late 2014 and continued regularly [25], but has never been implemented in Busia district.

### Entomological surveillance

At study initiation, 100 and 80 households were randomly selected and enrolled in the Nagongera cohort and Tororo-Busia cohort, respectively. Household selection of both studies is described in detail elsewhere [23, 26]. Although the emphasis of this report lies in the entomological surveillance conducted in Tororo-Busia from 2020-2022 and in Nagongera from 2017-2018, we place our findings in context using previously published entomological surveillance data from Nagongera before 2017 [26].

Between 2011 and 2017, mosquitoes were collected on a monthly basis in all Nagongera households using CDC light traps (Model 512; John W. Hock Company, Gainesville, FL). Miniature CDC light traps (Model 512; John W. Hock Company, Gainesville, Florida, USA) were positioned with the light 1 m above floor level at the foot end of a bed. Traps were set in all bedrooms at 7:00 PM and collected at 7:00 AM the next morning. From 2017 to 2018, 80 households from Nagongera were re-enrolled and collections were intensified to a biweekly timeframe. To enhance collection of blood fed mosquitoes, resting and exit trap collections were conducted every two weeks during the two rainy seasons in Nagongera from 2017 to 2018. Resting mosquitoes were collected in all household bedrooms between 7 and 9 am by standard resting catches using Prokopack aspirators (Model 1419; John W. Hock Company, Gainesville, Florida, USA). Window exit traps (Muirhead-Thomson type) were set at 6:00 PM and were positioned over a window in bedrooms to catch any escaping mosquitoes. Any trapped mosquitoes were collected the following morning between 06:00 and 07:00 AM. In Tororo-Busia households, mosquitoes were collected every 2 weeks using CDC light traps as mentioned above. All trapped mosquitoes were stored individually for further processing.

Female *Anopheles* mosquitoes were assessed morphologically to determine taxonomy at the species level. Presence of sporozoites was determined using a standardized ELISA assay as previously described [23, 25]. Abdomens of all blood fed mosquitoes were separated from the thorax using dissecting pins and squeezed on filter paper (Whatman filter paper G23) and left to dry.

### Household selection for mosquito blood meal analysis

For efficiency reasons – avoiding the collection of non-informative human DNA profiles – households were selected for mosquito blood meal analysis based on the number of blood fed mosquitoes collected during the study period. Households from Nagongera or Tororo-Busia were selected if at least 5 or 9 blood fed mosquitoes were collected during the study period, respectively. This number was arbitrary but avoided resource intensive analyses on households with very few mosquitoes relative to the number of household occupants. To obtain human DNA for matching mosquito blood meals to cohort participants, one filter paper sample per participant was obtained for all study participants enrolled in selected households at the time of blood fed mosquito collections (2017-2018 for Nagongera households, 2020-2022 for Tororo-Busia households).

### Follow-up of study participants

All cohort study participants were provided access to an LLIN at enrolment and at enrolment, all participants reported their age and sex (female / male) assigned at birth (from now on ‘gender’). Follow-up of participants in Nagongera households consisted of unscheduled visits for all medical needs at a dedicated study clinic open 7 days a week plus scheduled visits every 3 months until September 2017 and every 4 weeks between October 2017 and October 2019 [23, 26]. Participants of Tororo-Busia households were followed-up using the same procedure with scheduled visits every 4 weeks. During scheduled visits or when presenting to the clinic with malaria symptoms, blood was collected for the assessment of parasitaemia by microscopy. *Plasmodium falciparum* parasites were also quantified molecularly during scheduled visits using the *var* gene acidic terminal sequence (varATS) quantitative polymerase chain reaction (qPCR), with a lower limit of detection of 0.05 parasites/µL using 200 µL whole-blood samples [27].

### Laboratory assays

To match mosquito bloodmeals with study participants, nucleic acids (NA) were extracted from filter papers of cohort participants and blood fed mosquito abdomens using automated extraction (MagNAPure 96 DNA and Viral NA Small Volume Kit – Roche Applied Science). Parasite biomass was quantified in total NA extracted from mosquito blood meals using quantitative PCR (qPCR) targeting the *Plasmodium falciparum* 18S rRNA gene. Briefly, 5uL of template was run in 15uL volume of TaqMan mix with a primer and probe concentration of 90 and 110 nM, respectively. Molecular analysis of extracted NA from both human and mosquito samples consisted of an Identifier PCR kit (AmpFLSTR Identifiler PCR Amplification kit, ThermoFisher, catalognr 4322288) targeting 15 microsatellite loci (Supplementary Table 1) and one sex-determining locus (Amelogenin). 1uL of template was run in 10uL final volume of Identifiler mix for 11 minutes at 95°C, for 28 cycles of 1 minute at 94°C, 59°C, and 72°C, followed by 45 minutes at 60 °C. Products were analysed a DNA analyser (Applied Biosystems 3730xl, 50 cm capillary length) with a 500 internal lane standard to determine size of the products. Alleles were called using GeneMarker V2.6.7 with a 100 relative fluorescence unit (RFU) threshold.

### Matching of STR profiles from mosquito bloodmeals and study participants

Population allele frequencies were calculated among human STR profiles from both cohorts. We used the bistro function with default parameters [22] in addition to a peak detection threshold of 100 RFUs. To match mosquito bloodmeals with cohort participants, pairwise bloodmeal-human log10LRs were used that compare the likelihood that the blood meal STR profile is from a certain cohort participant versus a random contributor. The two cohorts were analysed separately and the initial likelihood threshold was set at 1.5; in order to be a potential match the likelihood had to be above 30 times greater than that of a random contributor [22]. In addition to this first threshold, another dynamic threshold per blood meal was set based on the maximum log10LR of that blood meal (Supplementary Figure 1). This threshold was dynamic in the sense that when the number of matches was less than the number of estimated contributors (NOC) this threshold was decremented by 0.5 to find a number of matches that is above or equal to the NOC. The match set was considered with highest confidence when two values of the dynamic threshold were met and the number of bloodmeal-human pairs was maximised. If no match sets were identified by a dynamic threshold of 1, or when all match sets had more than NOC pairs, the algorithm returned no bloodmeal-human pairs.

### Statistical analysis

To assess factors related to the number of nightly bites, we fitted a multilevel negative binomial model using the glmmTMB package in R. The response variable was set as the number of blood fed mosquitoes matched to a cohort participant per trapping night. We included participants who reported to have slept in the household during a trapping night in which at least one mosquito was successfully STR profiled. The model assumed a negative binomial distribution for the response variable and a random person-effect was included in the model to account for within-individual correlation. The model accounted for several adjustors (Appendix methods) and the following variables were included as potential risk factors: participant age (in categories), participant gender, whether a participant was reported to have slept under an insecticide treated net during the night of mosquito collection, and whether a participant was infected with *Plasmodium falciparum*. The latter was defined by a nearest microscopy slide reading between -28 and +7 days from the trapping night (Appendix methods); this time-window rules out the possibility that the blood-stage infection was a consequence of the observed mosquito bite. Sensitivity analyses were performed on the time-window used for the *P. falciparum* infection risk factor (Appendix methods).

To study biting biases in our set of mosquito-human matches, we used the Gini index as a measure of statistical dispersion of nightly bites, and 95% Confidence Interval (CI) by using the Gini function in the R package DescTools v0.99.54. The average nightly bites per study participant was used as input and calculated as the sum of bloodmeals matched to the individual divided by the trapping person-nights in the study. A random distribution in mosquito bites was defined by every individual having the same probability of being bitten, whereas an even distribution of bites was defined by every individual receiving the same number of mosquito bites. We computed the Gini index under a random distribution of bites by simulating 1000 bites with a random distribution where the probability of being bitten was weighted by the number of person-trapping nights in the study.

Differences in proportions were assessed using a z-test. All p-values below 0.05 were considered significant. Data analysis was performed in R (v2022-03-10).

### Ethical approval

Written informed consent was received from all participants prior to enrolment. Ethical approval for both cohort studies was obtained from the Makerere University School of Medicine Research and Ethics Committee, the Uganda National Council of Science and Technology, the London School of Hygiene & Tropical Medicine Ethics Committee, and the University of California, San Francisco.

## Results

### Indoor mosquito exposure

Between 2011-2019, a total of 147,989 female *Anopheles* mosquitoes were collected in Nagongera households in 9,230 overnight CDC light trap collections. Between 2017-2018, a total of 2,240 overnight exit trap collections and 2,239 resting catches resulted in trapping an additional 212 and 558 *Anopheles* mosquitoes, respectively. The median number of *Anopheles* collected in CDC light traps decreased from 30.3 (interquartile range (IQR) 20.6-45.0) per household trapping night between 2011-2014 to 3.33 (IQR 1.11 – 4.56) between 2015-2016 and 2.65 (IQR 1.75-4.31) between 2017-2018. Of all mosquitoes collected between 2011-2019, 90.1% (133,365/147,989) belonged to the *Anopheles gambiae s.l.* species complex.

In the Busia-Tororo cohort, a total of 52,468 *Anopheles* mosquitoes were collected in 3,081 overnight CDC light trap collections between 2020 and 2022 in 80 households, 20 located in Busia and 60 in Tororo district. The median number of *Anopheles* collected per trapping night was 13.5 (IQR 6.84-26.6) per household in Busia district, compared to 15.2 (IQR 8.40-25.6) in Tororo households. The majority of mosquitoes collected in Tororo-Busia households between 2020-2022 belonged to the *Anopheles gambiae s.l.* (68.3%, 35,883/52,468) species complex, followed by *Anopheles funestus* mosquitoes (29.7%, 15,581/52,468).

### Collection of blood fed mosquitoes

For blood meal typing, 35 of 80 Nagongera households (43.7%) followed from 2017-2018 and 14 of 80 Tororo-Busia households (17.5%) followed from 2020-2022 were selected with at least 5 or 9 blood fed mosquitoes collected, respectively. Within the 49 selected households, a total of 20,848 *Anopheles* mosquitoes were collected using CDC light traps (Figure 1A). For Nagongera households, an additional 129 and 353 mosquitoes were collected in exit traps and resting catches, respectively. The number of mosquitoes caught per trapping night varied between households and was highest for households from Tororo-Busia (Figure 1B).

**Figure 1:**
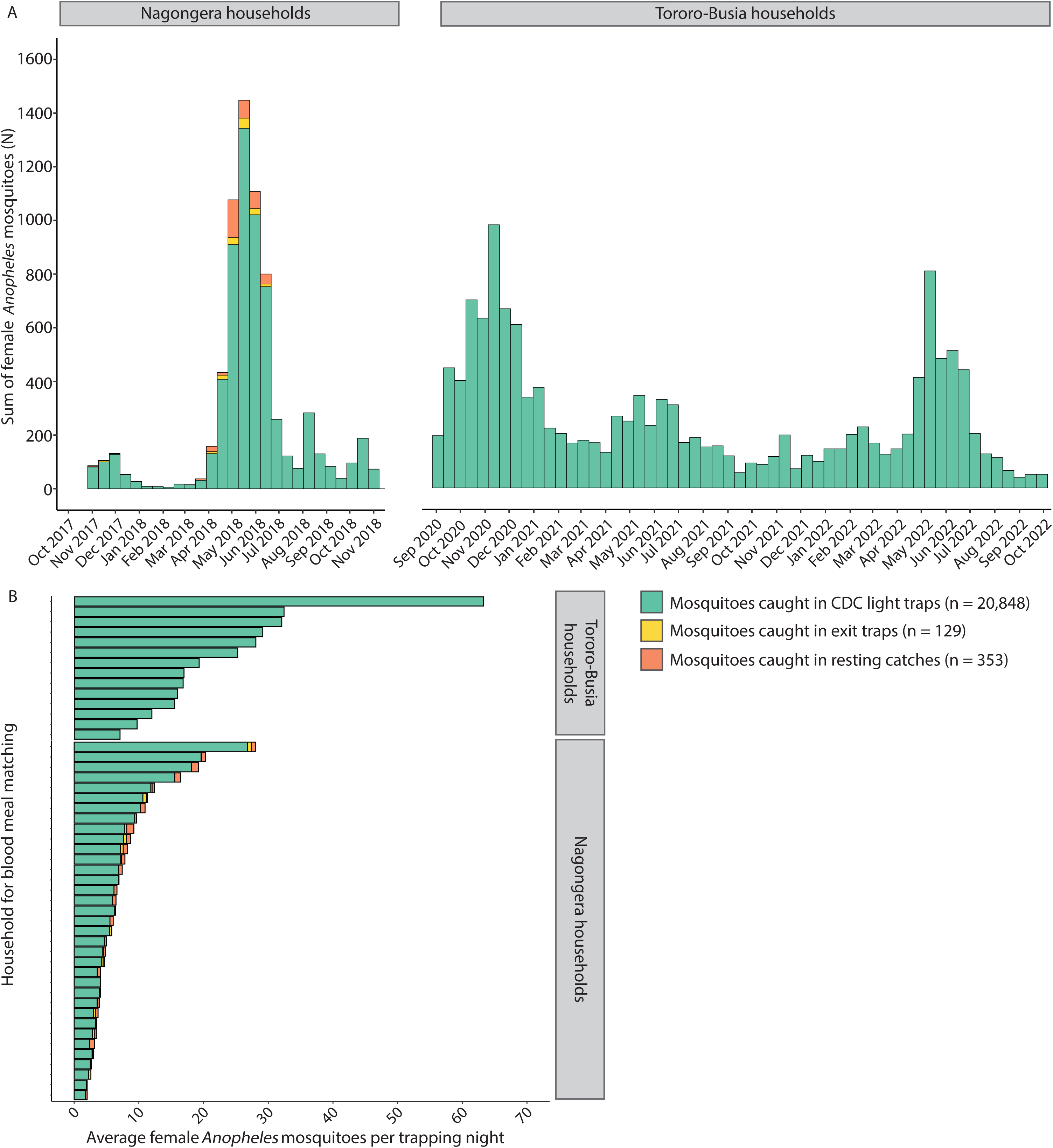
Female *Anopheles* mosquitoes collected in selected study households. Bar chart showing the number of female *Anopheles* mosquitoes caught over time (A) and the average number of mosquitoes caught per trapping night per household (B). A total of 35 and 14 households from Nagongera and Tororo-Busia were included, respectively. Colours indicate the trapping method used. Mosquito collections in Tororo-Busia households consisted of CDC-light traps only.

A total of 740 blood fed *Anopheles* mosquitoes were collected in the 49 selected households over time, of which 503 in Nagongera households between 2017-2018 and 237 in Tororo-Busia households between 2020-2022 (Figure 2A). From the blood fed mosquitoes collected in Nagongera, 86.2% (431/500) was collected using CDC light traps, 4.2% (21/500) using exit traps, and 9.6% (48/500) through resting collections; trap-level data was lost for 3 mosquitoes due to labelling issues. All 237 blood fed mosquitoes from Tororo-Busia households were collected using CDC light traps.

**Figure 2:**
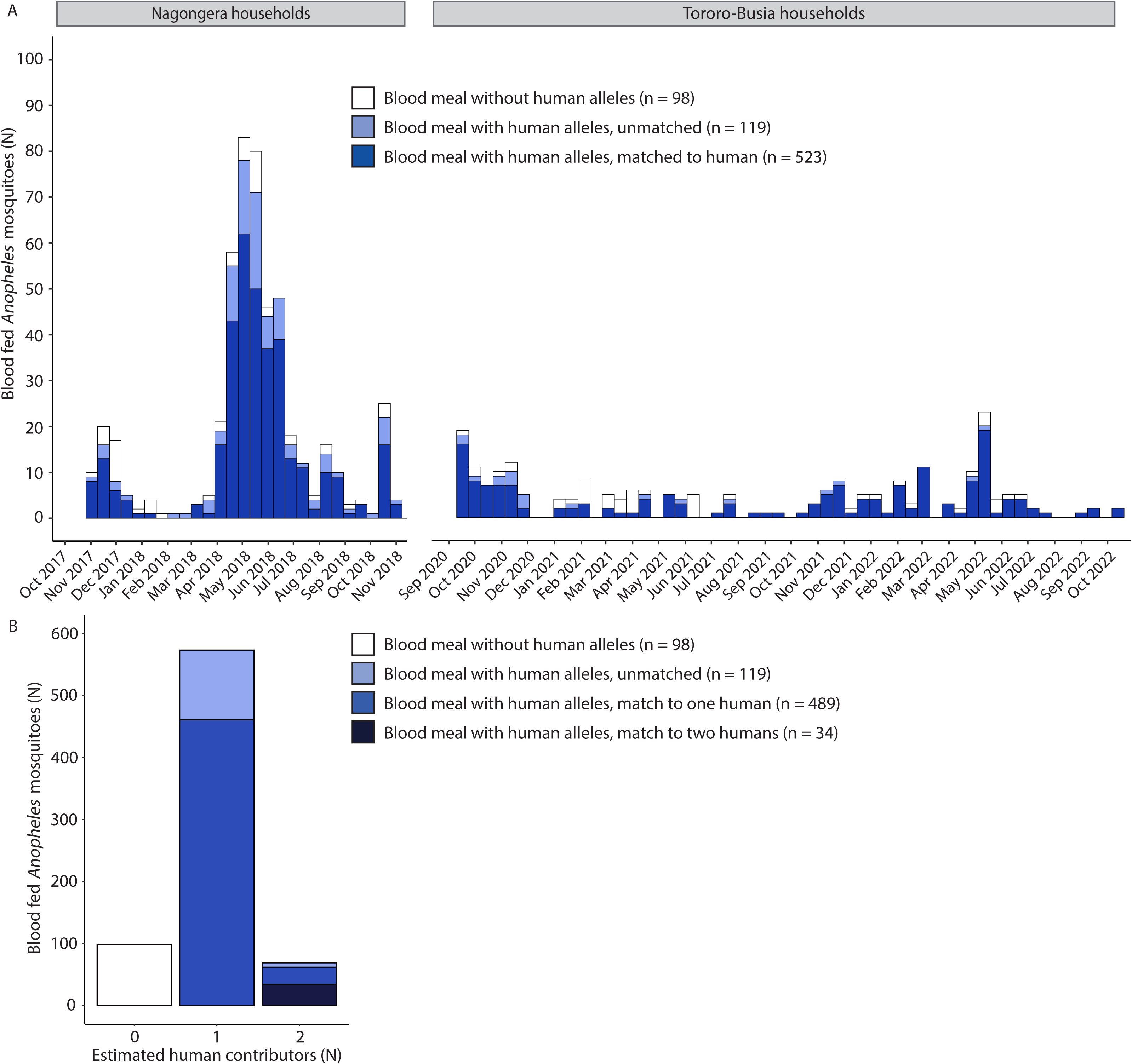
Blood fed *Anopheles* mosquitoes collected in selected study households. A) Bar chart showing the blood fed female *Anopheles* mosquitoes caught in 35 households from Nagongera and 14 households from Tororo-Busia over time. Colours indicate the status of STR typing, i.e. whether any human DNA was detected in the blood meal, and whether the identified human DNA was matched to a study participant. B) Bar chart indicating the number of blood fed *Anopheles* mosquitoes in relation to the estimated number of human contributors for both study sites combined. Based on the STR profile detected in mosquito blood meals, the number of human contributors was estimated. Colours indicate the status of STR typing, i.e. whether any human DNA was detected in the blood meal, whether the identified human DNA was matched to a single participant or to both human contributors for multi-sourced blood meals.

### High success rate in matching mosquito blood meals to one or two household residents

From 77.0% (570/740) of the collected mosquito bloodmeals, a complete DNA profile (i.e. 15 STR loci) could be obtained; 9.73% (72/740) and 13.2% (98/740) of bloodmeals had an incomplete (< 15 STR loci) or no human DNA profile, respectively (Figure 2B). Of the profiled (complete and incomplete) bloodmeals 10.7% (69/642) were estimated to have two human contributors, the remaining 89.3% (573/642) of bloodmeals was estimated to be single-sourced. The proportion of multi-sourced bloodmeals did not differ between Nagongera and Tororo-Busia households (P = 0.390) and no bloodmeals with more than two estimated human contributors were observed.

A complete STR profile was obtained for 98.7% (213/216) of the study participants enrolled in selected Nagongera households during mosquito collections. STR profiles of two participants were incomplete (14 STR loci) but still included in analysis, material of one participant did not result in an STR profile and was excluded from analysis. For Tororo-Busia households, we obtained STR profiles of 98.3% (114/116) of the study participants enrolled in selected households during mosquito collections. Material of two participants did not result in an interpretable STR profile; these participants were excluded from analysis. Two identical STR profiles were identified, labelled as a self-reported female and male participant that came from the same household. The Amelogenin sex-marker marked the two profiles as female; a filter paper from the female participant was likely labelled as the male participant and the male participant was excluded from further analysis. This resulted in the inclusion of 215 of 216 (99.5%) study participants from Nagongera households and 113 of 116 (97.4%) study participants from Tororo-Busia households in bloodmeal analysis (Supplementary Table 2).

The human source was identified for 80.2% (406/506) of complete mosquito bloodmeal profiles and 82.1% (55/67) of incomplete mosquito bloodmeal profiles that were estimated to have one human contributor. Both human contributors were found for 49.3% (34/69) of bloodmeals that were estimated to have two contributors based on their STR profile; 40.6% (28/69) was matched to one human contributor only with the other contributor likely coming from outside the sampled human population (Figure 2B). Amongst the matched blood fed mosquitoes, 99.6% (521/523) matched to a member from the household of collection. Five mosquitoes collected in Nagongera households matched a study participant from a participating household other than the one of collection which was located at 0.1-9 km distance, three of those were multi-sourced blood meals and also matched an individual from the household of collection.

### Parasite prevalence patterns in mosquito blood meals are comparable to those in humans

We next compared parasite prevalence in mosquito blood meals in the context of age-dependent parasite carriage as determined in cross-sectional surveys. In the human population, mean parasite prevalence in children under 5, children 5-15 years of age, and older individuals was 8.26%, 20.6% and 12.2% in Nagongera households, and 64.7%, 83.4%, and 60.7% in Tororo-Busia households, respectively. In matched blood fed mosquitoes, parasite prevalence patterns broadly mirrored those observed in the human population. In Nagongera households, parasite prevalence in mosquitoes was 8.33% (5/60), 13.4% (21/157), 17.0% (19/112) for children below 5, children 5-15 years of age, and older individuals, respectively. In Tororo-Busia, parasite prevalence in blood fed mosquitoes whose meal was linked to a child under 5 was 50.0% (15/30); parasite prevalence was 68.1% (47/69) and 41.9% (26/62) in blood meals linked to children 5-15 years of age and older individuals, respectively (Figure 3). Of the 228 blood-fed mosquitoes from Tororo-Busia households screened for sporozoite presence, only 1.31% (3/228) was positive. The overall sporozoite rate in female *Anopheles* mosquitoes collected between 2020-2022 and between 2017-2018 from all Tororo-Busia (n = 80) and Nagongera (n = 80) households was 1.17% (459/39,335) and 0.78% (8/10,234), respectively.

**Figure 3:**
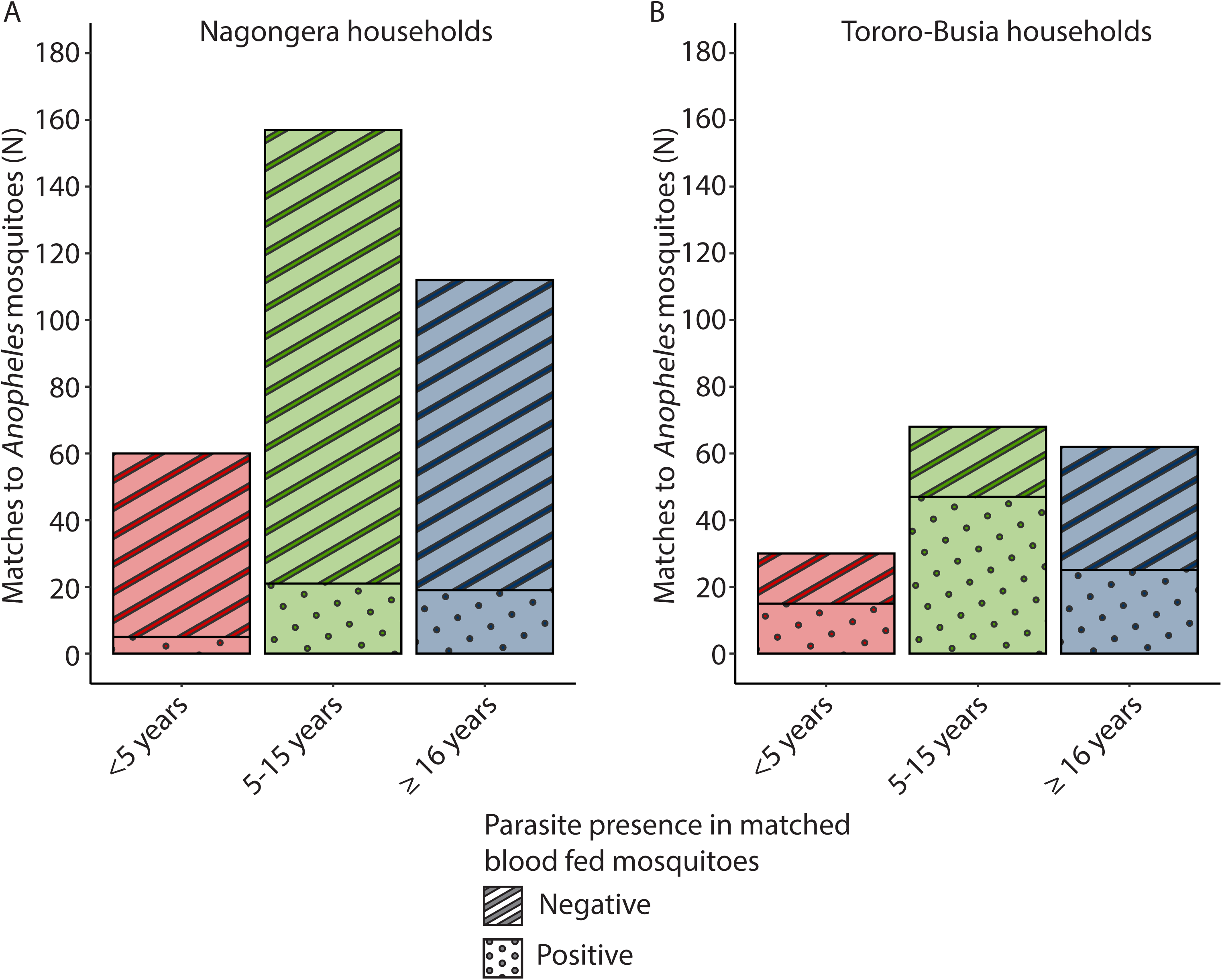
Parasite prevalence in matched blood fed mosquitoes in relation to participant age. Bar chart showing the proportion of matches to *Plasmodium falciparum* positive mosquitoes separated by age (in categories) for Nagongera households (A) and Tororo-Busia households (B). Bar colour indicates the age category and pattern reflects the infection status of the bloodmeal, being *P. falciparum* parasite positive or negative. Matched multi-sourced mosquitoes were excluded (n = 34) since blood was linked to individuals from multiple categories, leaving uncertainty to where the parasites originated from.

### Mosquito biting is heterogeneous

Of the participants from Nagongera households, 39.5% (85/215) matched to more than one blood fed mosquito; 38.6% (83/215) did not match any mosquito, indicating they were not bitten by the blood fed mosquitoes collected. These proportions were similar in Tororo-Busia households, with 34.5% (39/113) and 41.6% (47/113) of participants matching to more than one or to none of the blood fed mosquitoes collected, respectively. The maximum number of blood fed mosquitoes matched to one participant across the study period was 18 and the nightly biting rate (i.e. the number of mosquito matches per trapping night) differed between study participants and between households (Figure 4A). The distribution of nightly bites was more heterogeneous (Gini index: 0.67, 95% CI: 0.63-0.71) than expected by random chance (Gini index: 0.46, 95% CI: 0.43-0.51), with 20.0% of study participants receiving 67.0% of the observed mosquito bites per trapping night (Figure 4B) without an apparent difference between study sites (Supplementary Figure 2).

**Figure 4:**
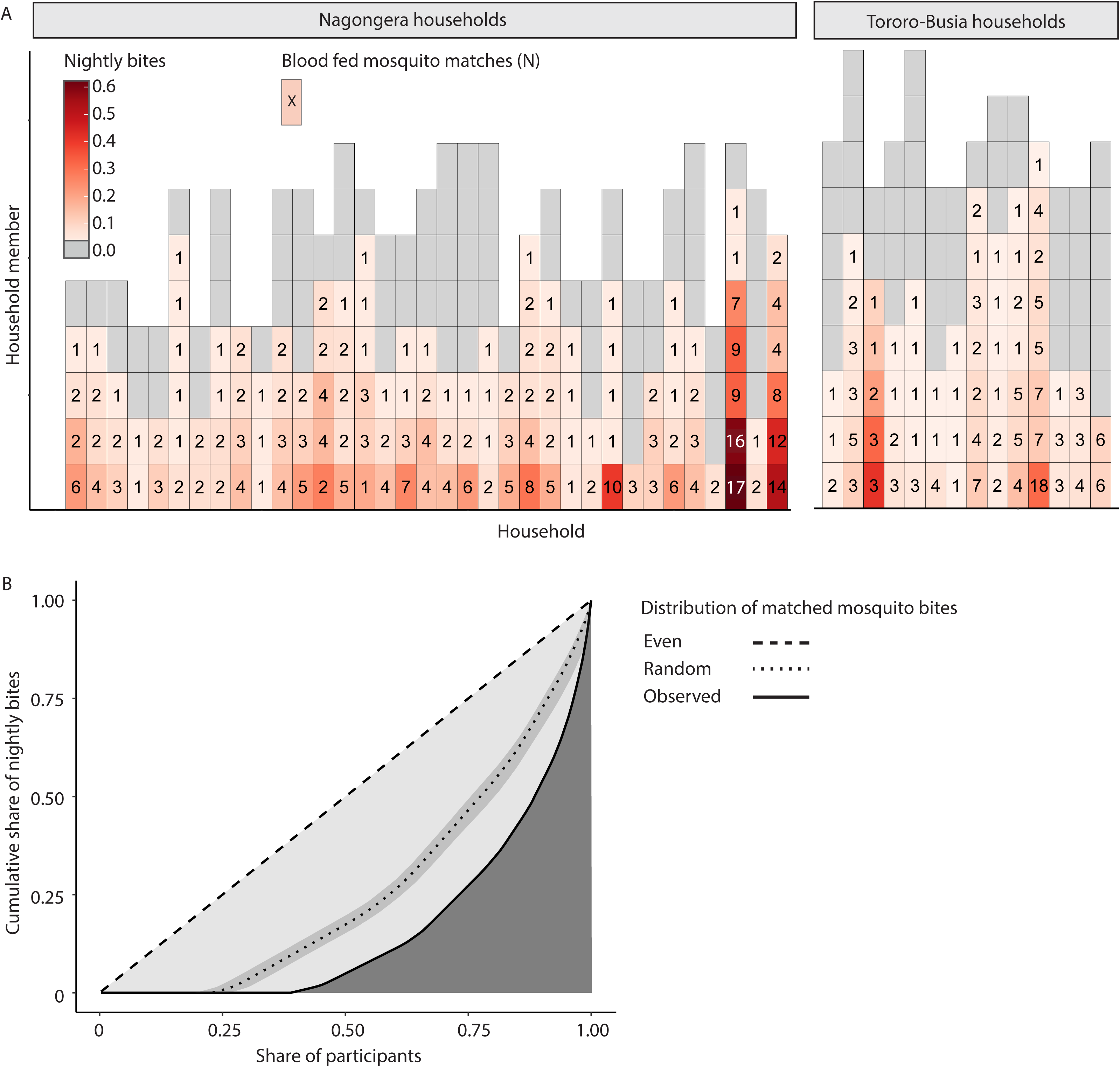
Heterogeneity in mosquito biting. A) Heatmap showing the variation in nightly bites (i.e. bites per trapping night) and the absolute mosquito matches. Rectangles indicate individual participants, participants were organized by household on the x-axis. Households were organized by the total number of *Anopheles* mosquitoes collected during the study period in ascending order for both study sites separately. Colours reflect the individual nightly bites given by dividing the total number of mosquito bites by the number of experienced trapping nights. The numbers indicated in the rectangles show the absolute number of blood fed mosquitoes matched to the individual across the study period. B) Distribution of nightly bites. Lorenz curve showing the share of participants versus the cumulative share of nightly bites for both study sites combined. The lines indicate the share under an even (dashed line), random (dotted line) and the observed (solid line) distribution. Under a random distribution, simulated by 1000 bite distributions across the number of study participants (n = 324) with known trapping night data, the Gini index was (0.44, 95% CI: 0.41-0.48). Under the observed distribution, the Gini index was 0.67 (95% CI 0.63-0.72), demonstrating higher heterogeneity than expected under a random distribution.

### Variation in mosquito biting is consistently related to human age

To better understand drivers of heterogeneity in nightly bites, we investigated participant population demographics in relation to mosquito biting. Whilst taking into account the number of household occupants present during trapping nights, children below the age of 5 years accounted for the lowest proportion of nightly bites (19.9%), followed by children aged 5-15 years (42.2%) and individuals aged 16 years or older (37.9%, Figure 5A). The received bites per trapping night was significantly lower in children aged below 5 years compared to the two older age groups (P < 0.0001); the difference between children aged 5-15 years and individuals aged 16 years or above was not statistically significant (P = 0.25). The proportion of nightly bites for female participants was 43.3% compared to 56.7% for male participants, and the received number of bites per trapping night was significantly lower for female participants (P = 0.001, Figure 5A). Gender-related differences in mosquito exposure were greatest in Tororo-Busia households where male individuals aged 16 years or above were responsible for 29.1% of the observed bites, compared to 13.1% for female individuals of the same age group.

**Figure 5:**
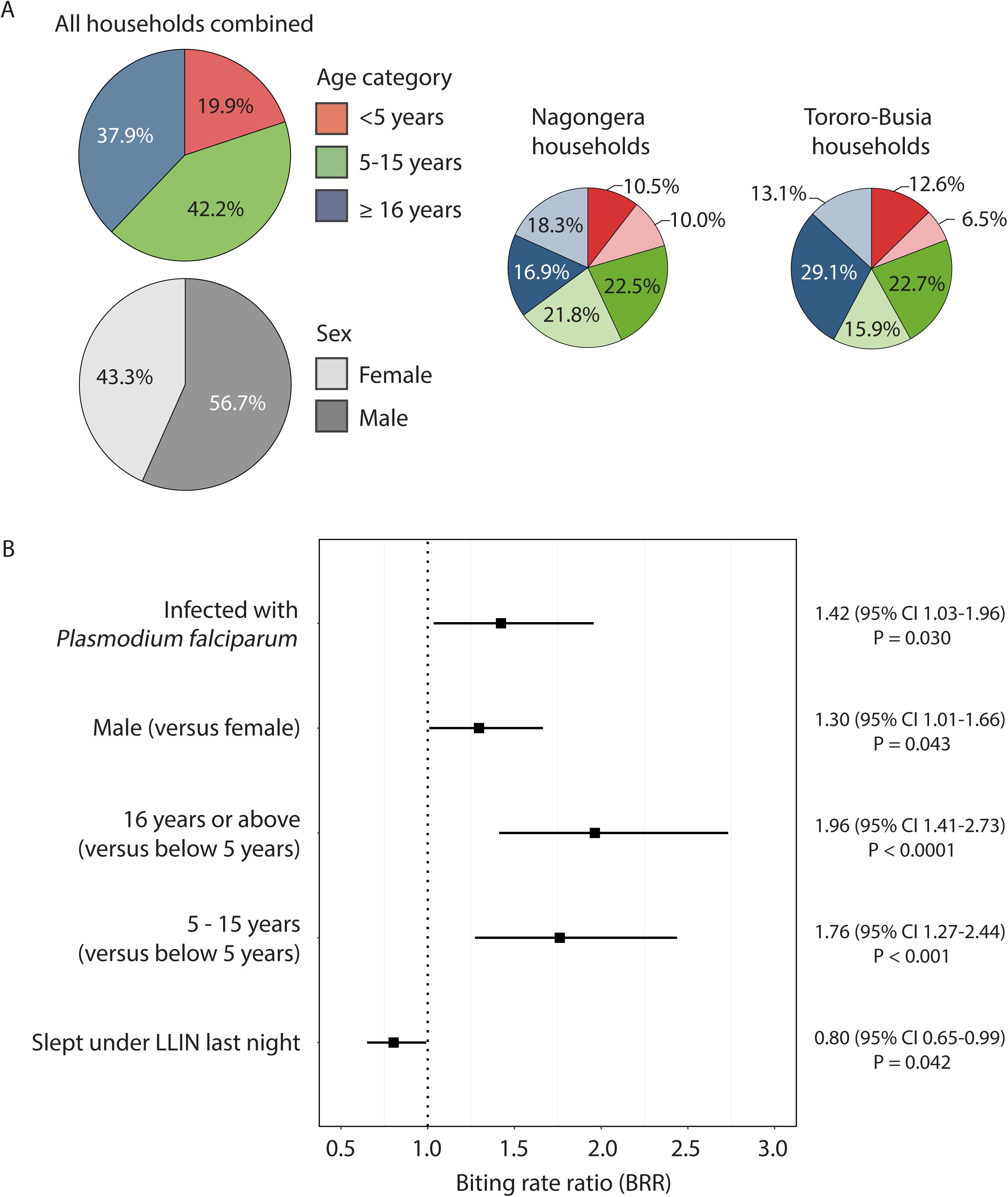
Mosquito biting in relation to participant demographics. A) Pie chart showing the proportion of nightly bites by participant age (in categories) and gender for both study sites combined for both sites separately. Age category is represented by colour and gender by shade. Size of pie chart segments represents the received number of bites per night at risk for a given category. For children aged below 5 years, the number of received bites was 106 during 3293 trapping nights, which differed from children aged 5-15 years (259/3790. P < 0.0001) and from individuals aged 16 years or above (191/3116, P < 0.0001). Female participants received 266 bites during 5562 nights at risk, which was less than the nightly bites received by male participants (290/4637, P = 0.001). B) Biting rate ratios for covariates in mosquito biting risk factor analysis. Covariates were assessed in multilevel risk factor analysis based on 1604 person-nights, selecting those nights with at least one STR-typed mosquito collected. Covariates included were: categorical age, gender, whether the participant reported to have slept under an LLIN the previous night, and whether the participant had a microscopy slide positive or negative for *P. falciparum* between 28 days prior and 7 days post trapping night. Adjustors included study site, number of STR-typed mosquitoes in the household that night, number of household members, and number of people in the sleeping space. Biting rate ratios are represented by squares, and 95% confidence intervals are depicted by lines.

We next used a multilevel approach to assess the effect of potential risk factors on the number of received mosquito bites. For this, data from trapping nights with at least one STR profiled mosquito was used and the effect of multiple predictor variables was assessed simultaneously. Combining data from both study settings, children aged 5-15 years had a mosquito biting rate ratio (BRR) that was 1.76 times that of children aged below 5 years (95% CI 1.27-2.44, P < 0.001, Figure 5B). The BRR of individuals aged 16 years or older was 1.96 times (95% CI 1.41-2.73, P < 0.0001) that of children aged below 5 years. Male participants had a BRR that was 1.30 that of female participants (95% CI 1.01-1.66, P = 0.030). The biting rate was lower in participants that reported to have slept under an LLIN during a trapping night (BRR 0.80, 95% CI 0.65-0.99, P = 0.042). Participants infected with *P. falciparum* had a BRR that was 1.42 times that of uninfected participants (95% CI 1.03-1.96, P = 0.030), although this effect did not remain statistically significant in sensitivity analyses using smaller time-windows over which parasite carriage in humans was determined (both P > 0.25, Supplementary Figure 3). The effects of participant age, gender, and net-usage remained statistically significant when removing the effect of *P. falciparum* infection (Supplementary Figure 4).

We next assessed the effect of LLIN usage in relation to participant age and gender. In a separate model with identical variables but with an interaction term between LLIN usage and participant age, we observed evidence of an interaction between LLIN usage and the oldest age category (≥ 16 years, P = 0.008). The biting rate of individuals aged 16 years or above who reported not using an LLIN the previous night was 2.85 times that of individuals aged below 5 years old (95% CI 1.86-4.37, P < 0.0001). This difference was no longer statistically significant (BRR 1.34, 95% CI 0.88-2.07, P = 0.176) when they did report LLIN use in the previous night. We did not observe an interaction between LLIN usage and the 5-15 year old age category (P = 0.529) or participant gender (P = 0.10). We neither observed evidence of an interaction between participant age, or participant gender, and study site (all P > 0.15).

## Conclusions

Mosquito biting is a non-random phenomenon and certain human individuals are bitten more than others. In vector borne diseases such as malaria, heterogeneity in mosquito biting may translate into heterogeneity in contributions to the human infectious reservoir. In the present study, we assessed heterogeneity in mosquito biting in relation to population demographics in two different endemicity settings. We observed that children below 5 years were bitten the least compared to school-age children (5-15 years) and adults (≥16 years). These patterns were consistent in two sites of different malaria endemicity.

Previous attempts to understand patterns in mosquito biting preference have mainly focused on matching human DNA in mosquito bloodmeals and were limited by the exclusion of partially digested and multi-sourced bloodmeals [5, 6]. In this study, we investigated mosquito biting heterogeneity using a novel statistical approach that allows for the inclusion of partially digested and multi-sourced bloodmeals [13, 22]. From all collected blood fed mosquitoes, 86.8% of blood meals were successfully typed as human blood meals of which 78.8% showed a complete STR profile from a single human that was linkable in 80.2% of the cases. Of the typed blood meals, 10.4% was single-sourced yet partially digested and 10.7% was observed to have multiple human blood sources. We successfully identified the human source for 82.1% of partially digested bloodmeals and we identified at least one human source for 89.9% of multi-sourced bloodmeals. By including partially digested and multi-sourced bloodmeals we increased our sample size of informative mosquito samples by 28.8%, thereby allowing a more accurate analysis of mosquito biting heterogeneity amongst different population groups.

Characterisation of multi-sourced bloodmeals can also have relevant implications for malaria transmission estimates [9]. If certain demographic populations are more likely to experience blood meals that are incomplete, for instance by increased uptake of interventions or defensive behaviour that results in interrupted/partial bloodmeals, ignoring multi-sourced bloodmeals can lead to biased biting estimates. In addition, entomological inoculation rate estimates are based on the sporozoite infection rate and human biting rate, which assumes that a mosquito takes a single blood meal per gonotrophic cycle. If mosquitoes take multiple blood meals and would expel sporozoites at each feeding attempt, this would increase number of infected bites a population is exposed to.

In our setting, young children (<5 years) were less frequently bitten by *Anopheles* mosquitoes than older individuals. This observed age pattern in *Anopheles* biting heterogeneity is in line with previous work in Malawi [6], Kenya [5, 13] and Papua New Guinea [14]. Previous studies in Thailand [11] and Puerto Rico [12] observed similar age patterns in bites of *Aedes aegypti* mosquitoes; adults were bitten more frequently than expected based on a random distribution. In addition to participant age, body surface area has been correlated with an elevated biting rate by *Aedes* [12] and *Anopheles* [4, 28] mosquitoes. Larger individuals with higher metabolic rates may release more CO_2_ and skin-volatiles that are important mosquito attractants [29, 30].

Besides host attractiveness, host availability can also influence mosquito biting heterogeneity. Adults experienced more mosquito bites than children in Tanzanian households with insecticide treated bed nets, an effect that was not observed in households without bed nets [8]. In our setting, the difference in biting rates between adults and young children was lower when adults reported to have slept under an insecticide-treated bed net the previous night. Even though adults are generally less likely to sleep under a bed net [31], bed net usage alone does not fully explain the observed age patterns in mosquito biting, since older individuals were also disproportionally bitten in absence [10, 12] or before implementation of bed nets [32]. This suggests that both uptake of protective interventions use and body size or other age-related attractants play a role in explaining differences in biting rates on different age groups.

Although differences in biting rates were also observed before implementation of bed nets [33], women are generally more likely to sleep under bed nets; this may potentially drive differences in mosquito biting between sexes [31]. In our setting, males were bitten more frequently than females, although we did not observe evidence of an interaction between bed net usage and participant gender. In contrast to the age pattern consistently observed in relation to mosquito biting, studies report conflicting results when assessing participant gender. In some settings, male participants were disproportionally bitten compared to female participants [2, 6, 13, 14, 33] whilst in other settings there was no difference observed [5, 9, 12]. Variations in mosquito exposure between sexes are likely (partially) driven by behavioural differences that are in turn culturally determined, making site-to-site variation inevitable. These behavioural differences may include indoor/outdoor activities around the household and bed times. It is also possible that differences in skin volatiles that exist between sexes [34] play a role.

Recent work from Kenya [13] reported higher biting rates in *P. falciparum* infected human individuals compared to uninfected individuals. Using a similar approach, we observed a comparable effect in participants with a microscopically detected *P. falciparum* infection -28 or +7 days from the trapping night. This could suggest that mosquitoes have a preference to feed on infected people; it is however not possible to rule out that other factors – for example related to behaviour – influence both risk of blood-stage infection and mosquito biting. Relevantly, the association between mosquito biting and human infection status did not survive sensitivity analyses in our study. Importantly, our study was not designed to collect household member and entomology samples concurrently, which would be required to assess the effect of a real-time *P. falciparum* infection on mosquito biting rates. Future studies on mosquito biting heterogeneity could therefore benefit from real-time data on human infection, ideally also including gametocyte presence. Moreover, the work from Kenya also showed that infectious mosquitoes bite infected human individuals [13]. Our study was not designed to evaluate this, since mosquito level sporozoite data was not available for Nagongera collections and sporozoite positivity was very low for Tororo-Busia collections.

Our study had several additional limitations. Firstly, we collected indoor resting mosquitoes only. Risk factors identified for mosquito biting rates may differ for mosquitoes that bite outdoors, potentially related to differences in host availability. Importantly, previous work in Papua New Guinea used outdoor barrier screens to catch blood fed *Anopheles* mosquitoes and observed similar age and gender patterns [14]. In addition, it is very likely that we did not capture all mosquito exposure; some mosquito exposure will have occurred outside sampling days and biting that did not result in a bloodmeal but can nevertheless involve sporozoite inoculation [35, 36], will have gone undetected. Nevertheless, we are confident that the mosquitoes collected resemble a random sample of the overall mosquito population and the undetected bites are likely to follow the patterns observed using the collected mosquitoes. Finally, blood samples were collected from individuals in participating households and we did no collect blood samples from neighbouring households. Including human blood samples from outside the study households could have increased the mosquito matching rates, which would have been interesting especially for multi-sourced blood meals.

In conclusion, we observed that adults and school-age children were disproportionally bitten by *Anopheles* mosquitoes in two malaria endemicity settings in Uganda. Previous work in the same setting identified school-age children as important contributors to the human infectious reservoir, followed by children below 5 years [27, 37]. When incorporating differences in mosquito exposure, older individuals, typically harbouring low-density infections, increase in relative importance for the infectious reservoir [4]. Our work further underscores the importance of school-age children in the human infectious reservoir.

## Captions of Artwork and Tables

**Supplementary Figure 1:** Matching algorithm used for identifying matches between short tandem repeat profiles form mosquito blood meals and household members. Log10LRs, log10 likelihood ratios; NOC, estimated number of human contributors; t, threshold. A detailed explanation and validation of the algorithm has been published elsewhere [22].

**Supplementary Figure 2:** Distribution of nightly bites by study site. Lorenz curve showing the share of participants versus the cumulative share of nightly bites for Nagongera households (A) and Tororo-Busia households (B). The lines indicate the share under an even (dashed line), random (dotted line) and the observed (solid line) distribution. Under a random distribution, simulated by 1000 bite distributions across the number of study participants (n = 215 for Nagongera households, n = 113 for Tororo-Busia households), Gini index for Nagongera households was 0.43 (95% CI 0.39-0.47) and 0.49 (95% CI 0.44-0.58) for Tororo-Busia households. Under the observed distribution, Gini index was 0.65 (95% CI 0.59-0.70) for Nagongera households, and 0.69 (95% 0.63-0.76) for Tororo-Busia households.

**Supplementary Figure 3:** Biting rate ratios for covariates in mosquito biting risk factor analysis. Biting rate ratios are represented by squares, and 95% confidence intervals are depicted by lines. Covariates were assessed in multilevel risk factor analysis based on 1604 person-nights. Covariates included were: categorical age, gender, whether the participant reported to have slept under an LLIN the previous night, and whether the participant had a microscopy slide positive or negative for *P. falciparum* between 7 days prior and 7 days post (A) or 14 days prior and 7 days post trapping night (B) Adjustors include study site, number of STR-typed mosquitoes in the household that night, number of household members, and number of people in the sleeping space.

**Supplementary Figure 4:** Biting rate ratios for covariates in mosquito biting risk factor analysis excluding infection status. Biting rate ratios are represented by squares, and 95% confidence intervals are depicted by lines. Covariates were assessed in multilevel risk factor analysis based on 1604 person-nights. Covariates included were: categorical age, gender and whether the participant reported to have slept under an LLIN the previous night. Adjustors included study site, number of STR-typed mosquitoes in the household that night, number of household members, and number of people in the sleeping space.

## Data Availability

All data produced in the present study are available upon reasonable request to the authors.

## Acknowledgements

We thank the study team and the Infectious Diseases Research Collaboration for administrative and technical support. We are grateful to the study participants who participated in this study and their families. We further acknowledge the helpful discussions with Christine Markwalter and Wendy O’Meara (Duke Global Health Institute, Duke University, US).

