## Appendix methods for "Anopheles mosquito exposure is associated with age, gender and bed net use in areas in Uganda experiencing varying malaria transmission intensity"

We used a multilevel model assuming a negative binomial distribution to assess potential risk factors in mosquito biting. We included data from individuals present in households during trapping nights in which at least one STR-typed mosquito was collected. The following variables were included as adjustors: the number of STR-typed mosquitoes collected in the household during a given trapping night, the study site (Nagongera versus Tororo-Busia), the number of household members enrolled in the study and the number of people sleeping in the same sleeping space as the person. We included as risk factors: participant sex, age (in categories of <5 years old, 5-15 years old and 16 years or older), whether the participant slept under a net during the trapping night, and whether the person was infected with *P. falciparum*. A participant was defined as infected or uninfected when, in the monthly survey, the nearest microscopy slide between -28 to +7 days of the trapping night was positive or negative, respectively. Observations were excluded if the participant had malaria and was treated up to 14 days before the trapping night, except when a subsequential microscopy slide reading confirmed the participant to be parasite negative. For sensitivity analyses, we restricted the definition of an infected (or uninfected) participant to the nearest microscopy slide reading between -14 to +7 days or -7 and +7 days of the trapping night.
