## Supplementary figures and images for "Anopheles mosquito exposure is associated with age, gender and bed net use in areas in Uganda experiencing varying malaria transmission intensity"

### Supplementary Figure 1

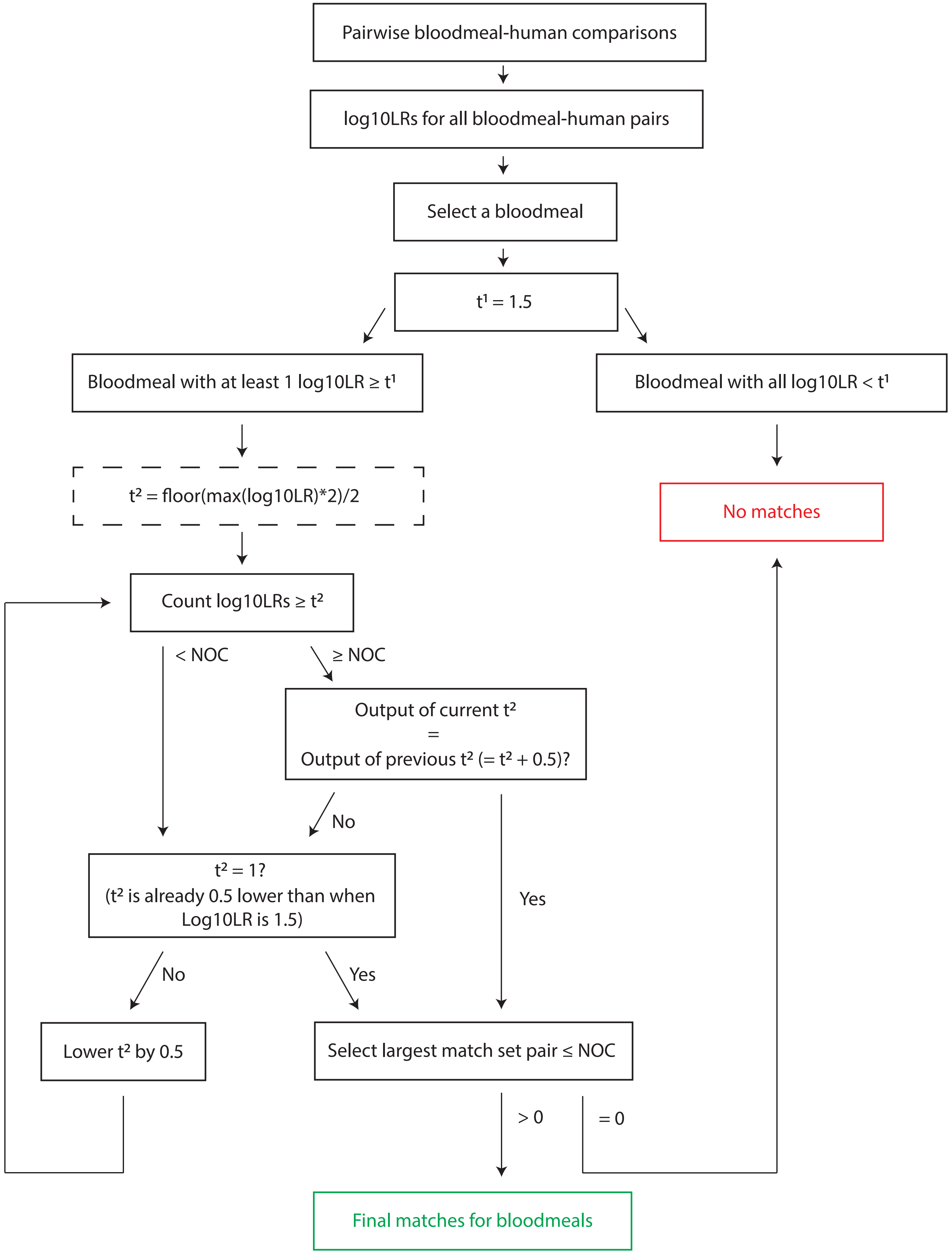

### Supplementary Figure 2

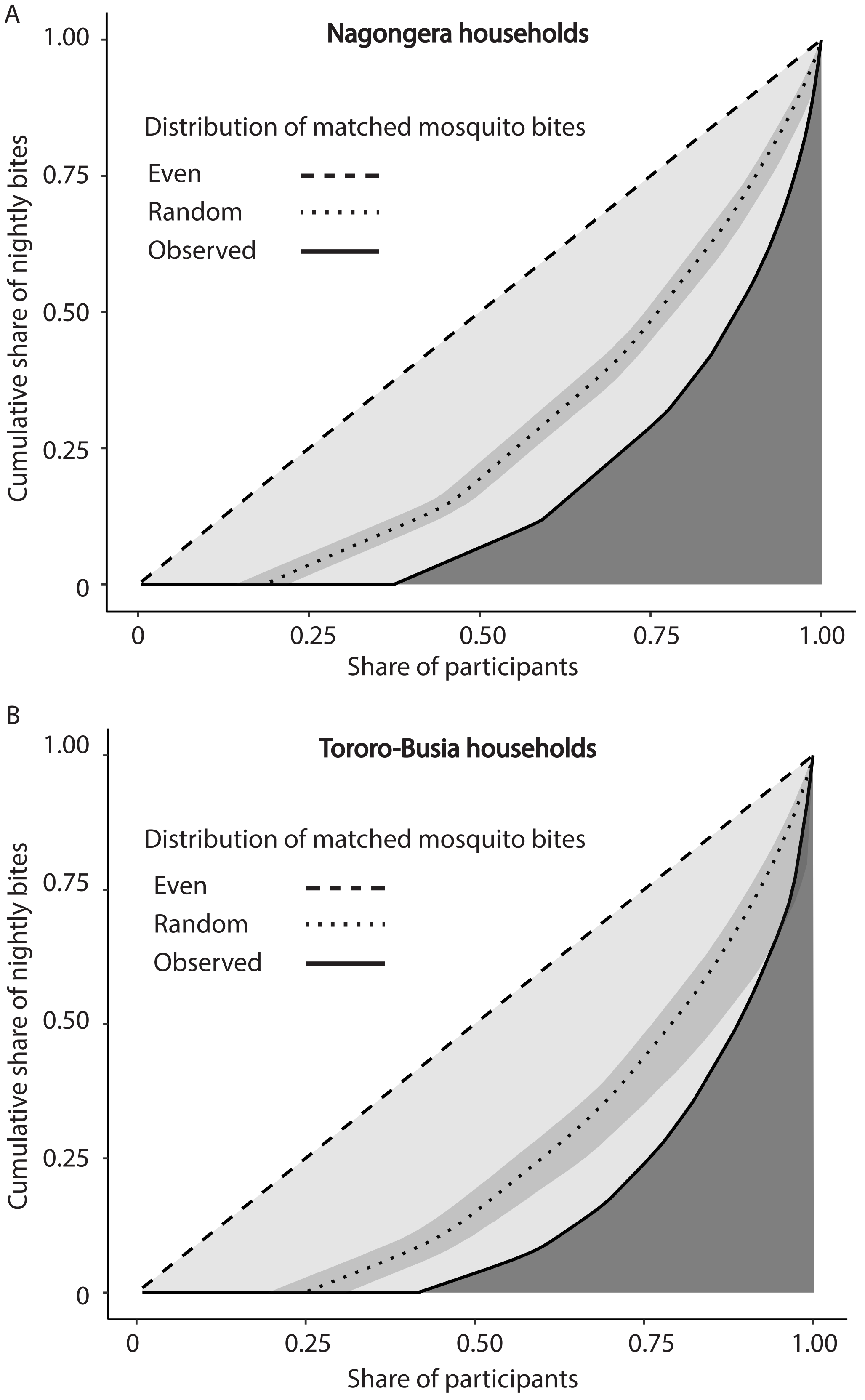

### Supplementary Figure 3

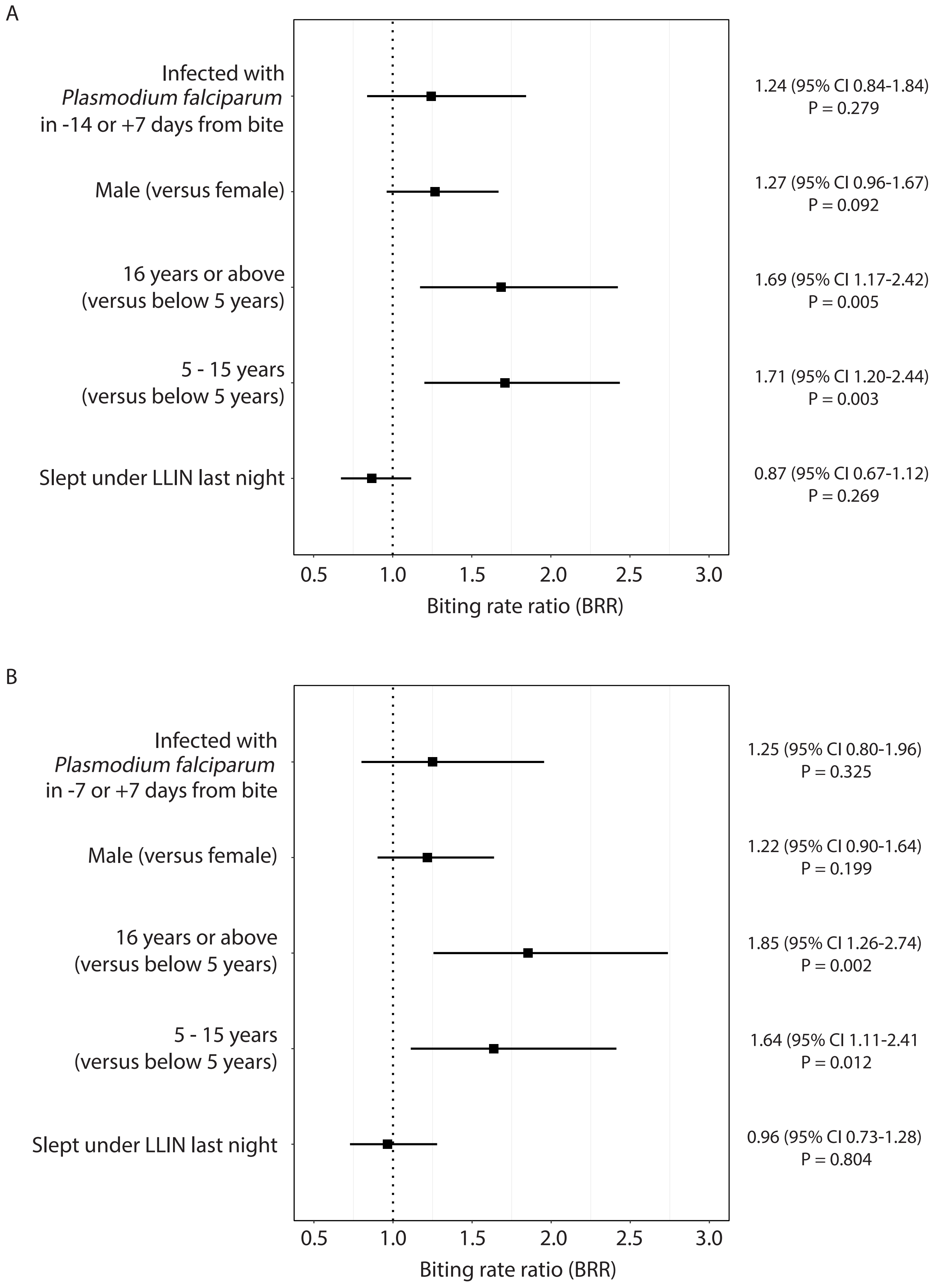

### Supplementary Figure 4

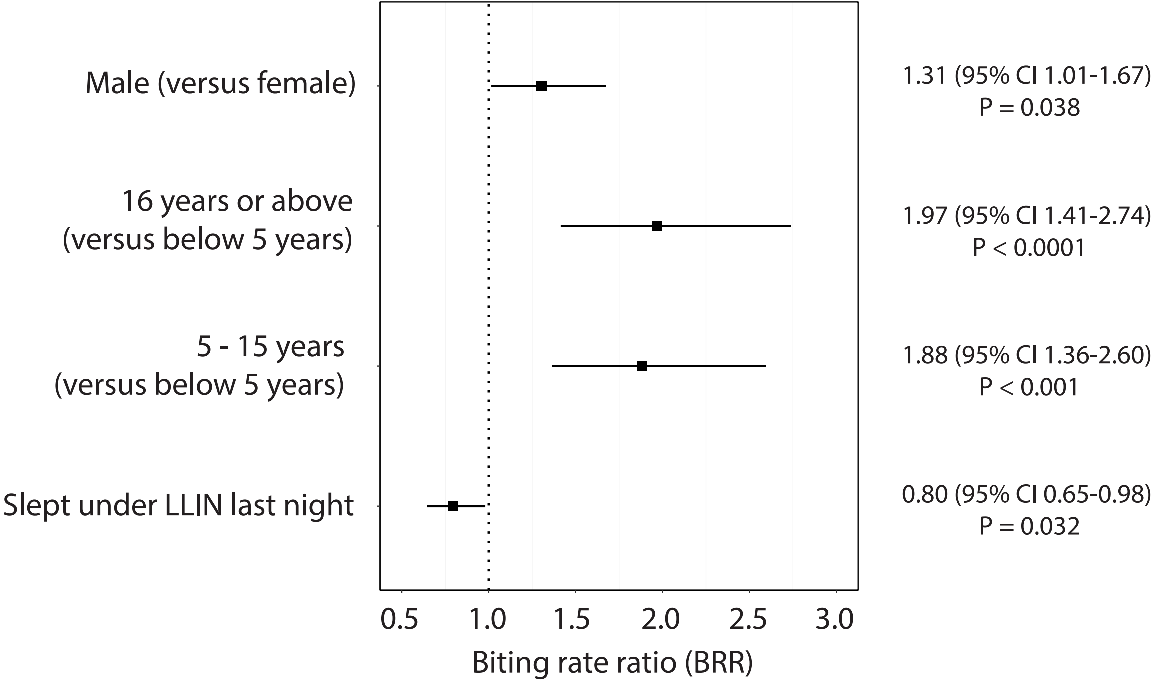
